# Comprehensive Serological Profile and Specificity of Maternal and Neonatal Cord Blood SARS CoV-2 Antibodies

**DOI:** 10.1101/2021.12.06.21267328

**Authors:** Rupsa C. Boelig, Sidhartha Chaudhury, Zubair H. Aghai, Emily Oliver, Francesca Manusco, Vincenzo Berghella, Elke Bergmann-Leitner

**Affiliations:** Department of Obstetrics and Gynecology, Division of Maternal Fetal Medicine, Sidney Kimmel Medical College, Thomas Jefferson University; Department of Clinical Pharmacology and Experimental Therapeutics, Sidney Kimmel Medical College, Thomas Jefferson University; Center for Enabling Capabilities, Walter Reed Army Institute of Research, Silver Spring, MD, USA; Division of Neonatology, Department of Pediatrics Nemours; Sidney Kimmel Medical College, Thomas Jefferson University; Department of Pathology, Sidney Kimmel Medical College, Thomas Jefferson University; Malaria Biologics Branch, Walter Reed Army Institute of Research, Silver Spring, MD

**Keywords:** COVID-19, pregnancy, passive immunity, immunity, serology

## Abstract

**Objective:** To describe the profile and specificity of maternal and neonatal cord-blood antibody profile in response SARS-CoV-2 virus exposure

**Methods:** This is a Prospective cohort study of delivering patients at Thomas Jefferson University Hospital from April 2020-February 2021. Primary objective was to describe unique maternal and fetal antibody epitope titers and specificity in those patients with COVID-19 history. Serologic profile assessed with a multiplex platform. Antigens used were: HA-trimer Influenza A (Hong Kong H3), spike trimers for SARS-CoV-2, SARS-CoV-1, MERS-CoV, and betacoronaviruses HKU-1 and OC43, as well as the spike N-terminal domain (NTD), spike receptor binding domain (RBD), and nucleocapsid protein (N; full length) for SARS-CoV-2.

**Results:** 112 maternal samples and 101 maternal and cord blood pairs were analyzed. Thirty-seven had a known history of COVID-19 (positive PCR test) in the pregnancy and of those, 17 (47%) were diagnosed with COVID-19 within 30 days of delivery. Fifteen of remaining seventy-six (20%) without a known diagnosis had positive maternal serology. For those with history of COVID-19 we identified robust IgG response in maternal blood to CoV2 nucleocapsid (N), spike (S) full-length and S (RBD) antigens with more modest responses to the S (NTD) antigen. By contrast, the maternal blood IgM response appeared more specific to S (full-length), than N, S (RBD) or S (NTD) epitopes. There were significantly higher maternal and cord blood IgG response not just to CoV2 spike (p < 10^−18^), but also CoV1 spike (p < 10^−9^) and MERS spike (p < 10^−8^). By contrast, maternal IgM responses were more specific to CoV2 (p < 10^−19^), but to a lesser degree for CoV1 (p < 10^−5^), and no significant differences for MERS. Maternal and cord-blood IgG were highly correlated for both S and N (R^2^ = 0.96 and 0.94).

**Conclusions:** Placental transfer is efficient, with robust N and S responses. Both nucleocapsid and spike antibody responses should be studied for a better understanding of COVID-19 immunity. IgG antibodies are cross reactive with related CoV-1 and MERS spike epitopes while IgM, which cannot cross placenta to provide neonatal passive immunity, is more SARS CoV-2 specific. Neonatal cord blood may have significantly different fine-specificity than maternal blood, despite the high efficiency of IgG transfer.

## Introduction

Pregnant women were found to be particularly vulnerable to respiratory pathogens and are more likely to need intensive care and have higher mortality due to SARS-CoV-2 infection^1 3^. SARS-CoV-2 infection (COVID-19) is associated with higher rates of pre-term birth^4^, pre-eclampsia^5,6^, and placental pathology ^7^.

Pregnancy significantly alters many elements of adaptive immunity in a gestational age-specific manner^8 10^. These adaptations have been studied in viral infection, e.g., in influenza A infection^11^. The effect of immunological adaptions in SARS-CoV2 infection during pregnancy has not been delineated. Initial studies on COVID-19 patients highlight several findings: CoV2-specific antibody (ab) responses are measurable 2-3 weeks after onset of symptoms^12,13^, and almost everyone who recovers from SARS-CoV2 infection develops ab responses^14^ that show a wide range of neutralization activity^15^. However, many questions remain such as the durability of these responses, the isotype profile at different stages of infection, and determinants of neonatal passive immunity. The presence/timing of neonatal immunity following maternal exposure can be a key consideration in repeat vaccination in pregnancy. As such, research into placental transfer of maternal antibodies and production of fetus-derived abs have aided in developing guidelines for timing pertussis and flu vaccinations for maximal maternal and neonatal benefit^16,17^. A study of serologic positive pregnant women found that IgG responses were higher in symptomatic vs. asymptomatic women, maternal IgM and IgG peaked 15-30 days post symptom onset, and passive IgG-immunity was found in about three-quarters of neonates and associated with maternal IgG levels^18^. Another study identified a high placental transfer ratio of antibodies, increased with latency between diagnosis and delivery^19^. These studies did not evaluate the range of SARS-CoV-2 antibody epitopes or cross reactivity with related viruses, limiting our understanding of the breadth of SARS CoV-2 antibody response as well as our ability to select optimal epitopes to follow in future research studies.

The objective of this study was to describe the profile and specificity of maternal serum and neonatal cord blood antibody response to maternal SARS-CoV-2 virus exposure

## Materials and Methods

This study was approved by Thomas Jefferson University Institutional Review Board as minimal risk, and each participant provided written consent. This study followed the Strengthening of Reporting of Observational Studies in Epidemiology (STROBE) reporting guidelines.

### Cohort Selection

This is a prospective cohort of pregnant patients consented for collection of samples at delivery, including maternal blood on admission and cord blood at delivery as part of an ongoing delivery cohort biorepository. Patients were consented either on admission for delivery or as an outpatient prior to planned delivery at our hospital. Per the study protocol additional maternal blood was collected on admission and cord blood collected at delivery. This cohort includes participants from April 2020 Feb 2021, however due to COVID related research restrictions, most of the participants were recruited November 2020-Feb 2021. Participants were included if they were consented and had maternal and/or cordblood samples available for this study. Those with prior COVID vaccination were excluded. Subjects were split into two groups-1) COVID-infected which was indicated by either documented SARS CoV-2 PCR test at any time in pregnancy *or* positive maternal spike IgG or IgM at delivery based on assays below or 2) COVID negative: no history of COVID-19 and negative SARS CoV 2 PCR testing on admission

### Data Collection

Electronic medical records were reviewed for demographic, medical, and obstetric history, date of first positive SARS-CoV-2 PCR test, date of delivery, antenatal complications, delivery outcomes. Latency between COVID-19 diagnosis and delivery was categorized as a binary outcome: within 30 days of delivery or >30 days from delivery. Standard of care at our institution includes universal SARS-CoV-2 PCR test on admission and neonatal SARS-CoV-2 PCR test if mother has a positive diagnosis on admission for delivery.

### Assays

#### Serological assessment

For assessing antibody specificity, a multiplex testing platform (Meso Scale Diagnostics, Rockville, MD): antigens manufactured in a mammalian expression system (Expi 293 F) are printed onto 10-plex plates. The antigens used were: HA-trimer Influenza A (Hong Kong H3), spike (soluble ectodomain with T4 trimerization domain) trimers for SARS-CoV-2, SARS-CoV-1, MERS-CoV, and betacoronaviruses HKU-1 and OC43, as well as the spike N-terminal domain (NTD, Q14-L303 of the SARS-CoV-2 spike sequence), receptor binding domain (RBD, R319-F541 of the SARS-CoV-2 spike sequence), and nucleocapsid protein (N; full length) for SARS-CoV-2, and bovine serum albumin (BSA) as negative control. Assays were performed blocked using Blocker A Solution and incubated at room temperature (RT) for 1h on a plate shaker, shaking at 700 rpm. The plates were washed three times with 1x MSD Wash Buffer. Sera were diluted to 1:1000 dilution with Diluent 100. Positive samples (pooled human serum from COVID-19 patients) and negative samples (pooled pre-pandemic human serum) were used as controls. Plates were sealed and incubated at RT for 2h on a plate shaker, shaking at 700 rpm, then washed three times with 1x MSD Wash Buffer. The detection antibody, SULFO-TAG either with anti-human IgG (or anti-human IgM antibody (was diluted to 2 μg/ml in Diluent 100 (MSD) and added to the wells and incubated at RT for 1h on a plate shaker. After washing, MSD GOLD Read Buffer B (was added to each well and immediately the plates were read on the MESO QuickPlex SQ 120 (MSD).

#### Placental histopathology

Placental histopathology was conducted as per routine clinical indications which includes COVID-19 at our institution. Clinical indications for sending placenta for pathology includes COVID-19 in pregnancy, maternal medical comorbidity such as hypertension or diabetes, antenatal complication such as chorioamnionitis, preterm delivery, nonreassuring fetal heart rate tracing. Placental findings were categorized as described in Supplemental Table 1.

### Outcomes

The primary outcome was to describe maternal antibody epitope profile and specificity related to COVID-19 exposure and correlate with cord blood levels. All antibody responses were analyzed following log transformation of the mean luminescence intensity read out of the MSD assay. Seropositivity for the CoV2 spike protein antigen was defined based on a cutoffs of 8.85 for IgM and 8.96 for IgG, in log transformed, as previously described^20^. Additional serological outcomes included: correlation between epitope levels, relation between antibody epitopes and latency to delivery

### Statistical Analysis

Statistical analysis conducted using SPSS v. 26.0 (Chicago, SPSS, Inc) and R. Continuous variables were compared with Mann-Whitney U test, categorical with Pearson Chi-Square analysis. Correlation between factors was assessed with bivariate correlation and reported with Pearson correlation coefficient. P<0.05 considered significant for all analyses. Figures were generated using the *stats, ggplot2*, and *corrplot* packages in R.

## Results

During the study period there were 112 maternal samples including 101 maternal and cord blood pairs collected. Thirty-six had a known history of COVID-19 (positive PCR test) in the pregnancy and of those, 17 (47%) were diagnosed with COVID-19 within 30 days of delivery. Fifteen of the remaining seventy-six without a known diagnosis had positive maternal serology (IgG or IgM to SARS-CoV-2 spike) positive serology; this was reflected in positive cordblood IgG as well. This represents a 20% seroprevalence rate among study participants.

### Baseline Characteristics

Baseline characteristics are described in Table 1. There were 51 in the COVID infected group (n=40 with maternal and cordblood paired sample available). Black and Hispanic patients were disproportionately represented in the COVID infected group. Severity of COVID illness/symptoms was documented for 32 of 36 known cases with the large majority (N=30) being asymptomatic or mild, and 2 being moderate severity.

**Table 1:** Comparison of Baseline Characteristics and Pregnancy Outcomes in Pregnant Women with and without PCR or serological evidence of COVID-19.

| Demographics | Maternal COVID PCR<br>and Serology<br>negative<br>(N=61) | Maternal COVID PCR<br>or Serology positive<br>(N=51) | P-value |
| --- | --- | --- | --- |
| Race |  |  | 0.03 |
| White (Non-Hispanic) | 35 (57) | 18 (35) |  |
| Black (Non-Hispanic) | 19 (31) | 23 (45) |  |
| Asian | 2 (3) | 0 (0) |  |
| Hispanic | 5 (8) | 10 (29) |  |
| Chronic Hypertension | 7 (12) | 8 (16) | 0.56 |
| Pregestational Diabetes | 0 | 2 (4) | 0.21 |
| Primiparous | 33 (54) | 30 (59) | 0.62 |
| Preterm birth (<37 weeks) | 4 (7) | 10 (20) | 0.04 |
| Preeclampsia | 16 (26) | 14 (28) | 0.88 |

### Maternal SARS-CoV-2 serology

Of the 36 with PCR confirmed COVID-19 infection, 92% had positive S-IgM (N=33) and 92% (N=33) had positive S-IgG. We found that subjects with reported COVID-19 diagnosis have high CoV2 S protein-specific IgM and IgG levels in maternal blood, and high IgG but low IgM levels in cord blood, consistent with the dogma that IgM does not cross the placenta (Figure 1). Among subjects with no prior COVID-19 diagnosis, 20% (15 of 76) were seropositive for CoV2 spike protein which is consistent with serosurveillance studies^21^.

**Figure 1:**
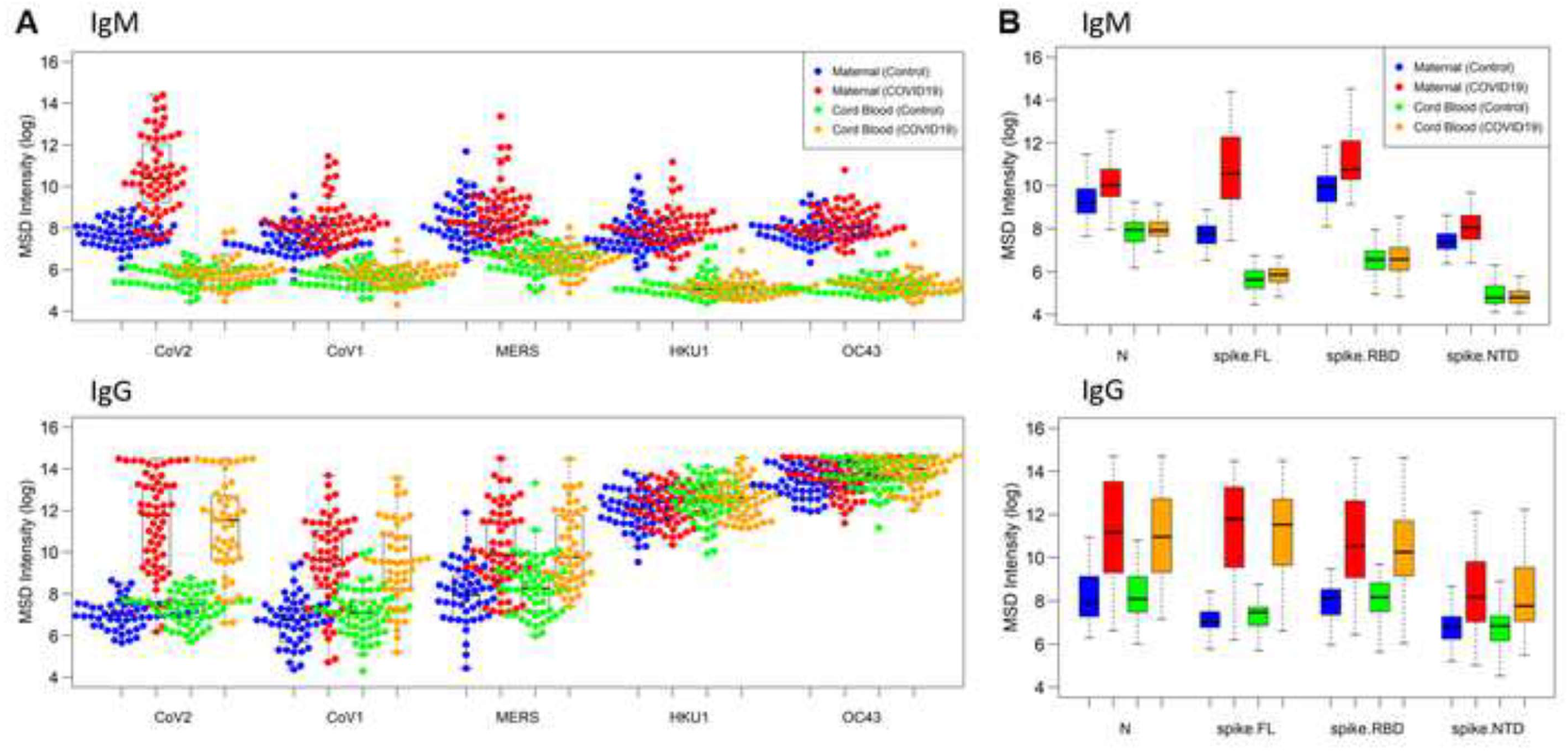
Antibody responses from maternal and cord blood samples collected from 91 pregnant women. Maternal and Cord Blood COVID-19 refers to samples from patients with known COVID-19 history (PCR confirmed) or positive maternal serology. Data reported as natural log-transformed luminescence signal. A) CoV-specific IgM responses (top) and CoV-specific IgG responses (bottom) in maternal sera and cordblood samples to the spike proteins of SARS-CoV1, SARS-CoV2, MERS-CoV, HKU-1, and OC43. Participants with prior COVID-19 history had significantly higher responses not just to CoV2 spike (p < 10^−18^), but also CoV1 spike (p < 10^−9^) and MERS spike (p < 10^−8^). By contrast, for IgM responses in maternal blood, participants with prior COVID-19 history showing significantly higher response to CoV2 (p < 10^−19^), but to a lesser degree for CoV1 (p < 10^−5^), and no significant differences for MERS. B) Fine specificity of SARS-CoV-2 specific IgM (top) and IgG ((bottom) responses in maternal sera and cordblood samples to SARS-CoV2 epitopes, i.e., nucleoprotein (N), the full-length spike protein (spike-FL) and its functional subdomains, i.e., receptor binding domain (RBD) and N-terminal domain (NTD). Subjects with prior COVID-19 infection had robust IgG response in maternal blood to CoV2 N, S (full-length) and S (RBD) antigens that were approximately 20-fold, 150-fold, and 10-fold higher, respectively, than what was found in subjects without prior infection, with more modest responses to the S (NTD) antigen. By contrast, maternal blood IgM response to S (full-length) was approximately 20-fold higher in COVID-19 subjects than in subjects with no prior COVID-19 infection history, but only 2-to 3-fold higher for N or S (RBD).

In terms of the magnitude and epitope specificity of the CoV2 antibody response, we found that subjects with prior COVID-19 infection had robust IgG response in maternal blood to CoV2 N, S (full-length) and S (RBD) antigens that were approximately 20-fold, 150-fold, and 10-fold higher, respectively, than what was found in subjects without prior infection, with more modest responses to the S (NTD) antigen (Figure 1B). By contrast, the maternal blood IgM response appeared more specific to S (full-length), than N, S (RBD) or S (NTD) epitopes. For example, maternal blood IgM response to S (full-length) was approximately 20-fold higher in COVID-19 subjects than in subjects with no prior COVID-19 infection history, but only 2-to 3-fold higher for N or S (RBD) (Figure 1B).

### SARS CoV-2 positive and relation cross reactivity to related viruses

We evaluated maternal IgG and IgM response to different coronavirus spike proteins to assess the cross-reactivity of these antibody responses across coronaviruses. For IgG responses on maternal blood and cordblood, we found that participants with prior COVID-19 history had significantly higher responses not just to CoV2 spike (p < 10^−18^), but also CoV1 spike (p < 10^−9^) and MERS spike (p < 10^−8^), suggesting a largely cross-reactive IgG response. By contrast, for IgM responses in maternal blood, we found the response more specific to CoV2, with participants with prior COVID-19 history showing significantly higher response to CoV2 (p < 10^−19^), but to a lesser degree for CoV1 (p < 10^−5^), and no significant differences for MERS (Figure 1A).

### Cordblood SARS CoV-2 Serology

Cordblood responses largely mirrored maternal blood responses for IgG with respect to magnitude and epitope specificity (Figure 1). As expected, cordblood IgM responses were approximately 50-to 400-fold lower than their corresponding IgM responses in maternal blood. Finally, principal component analysis (PCA) plot of IgG and IgM responses to CoV2 antigens in the panel shows that samples with prior COVID-19 exposure are clearly distinguishable from samples without prior exposure, and that maternal samples are clearly distinguishable from cordblood samples (Figure 2).

**Figure 2:**
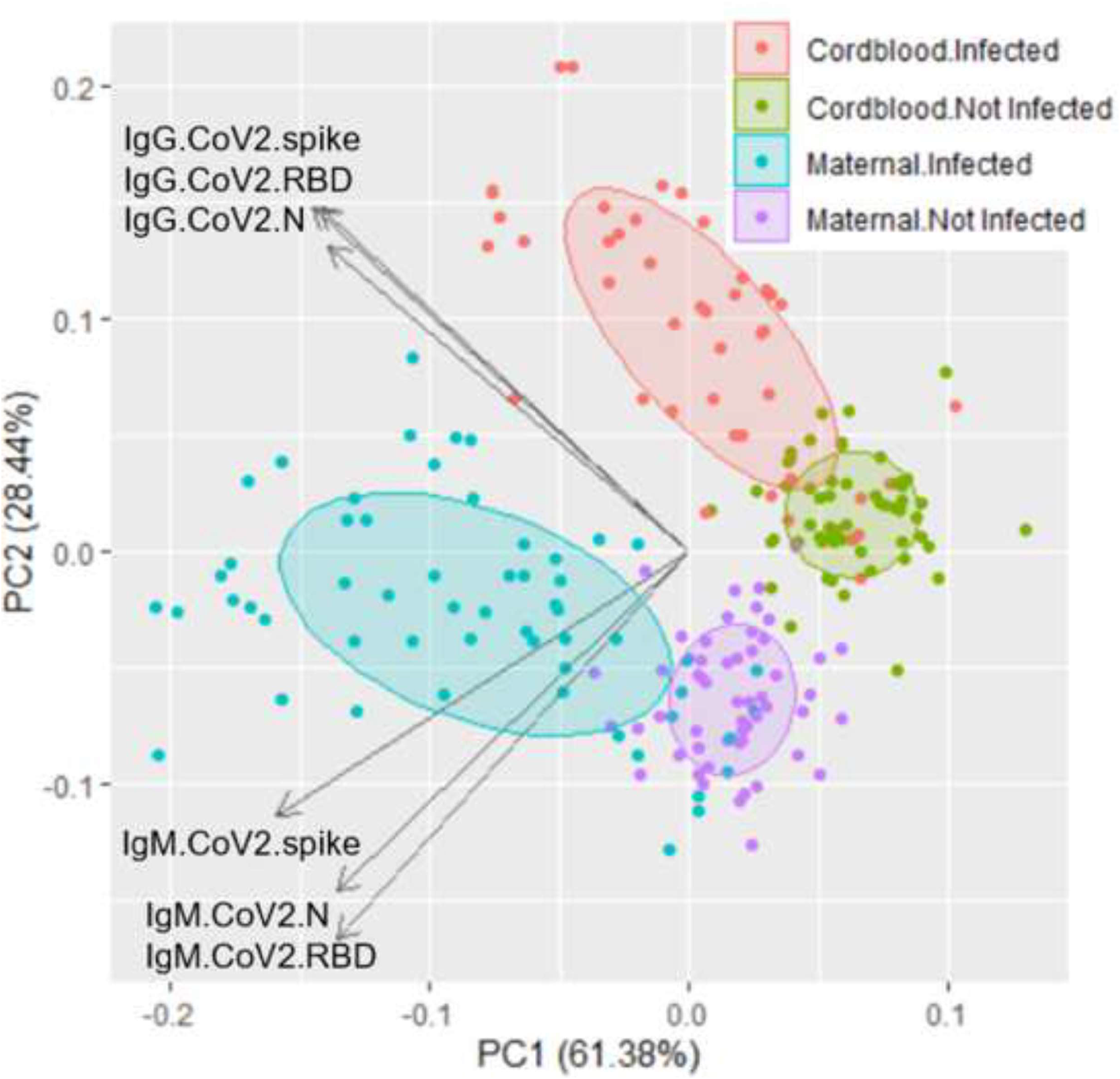
Principal Component Analysis of antibody responses. IgM and IgG responses to CoV2 antigens N, spike, and RBD clearly distinguish between prior COVID-19 cases and maternal from cord blood.

We found a high correlation of IgG (R^2^ = 0.96 and R^2^ = 0.94), but not IgM responses (R^2^ = 0.13 and R^2^ = 0.01) between paired maternal and cord blood samples for CoV2 spike and N antigens, respectively (Figure 3). A linear fit between maternal and cordblood IgG responses shows a slope of 1.01 for both CoV2 spike and CoV2 N antigens respectively.

**Figure 3:**
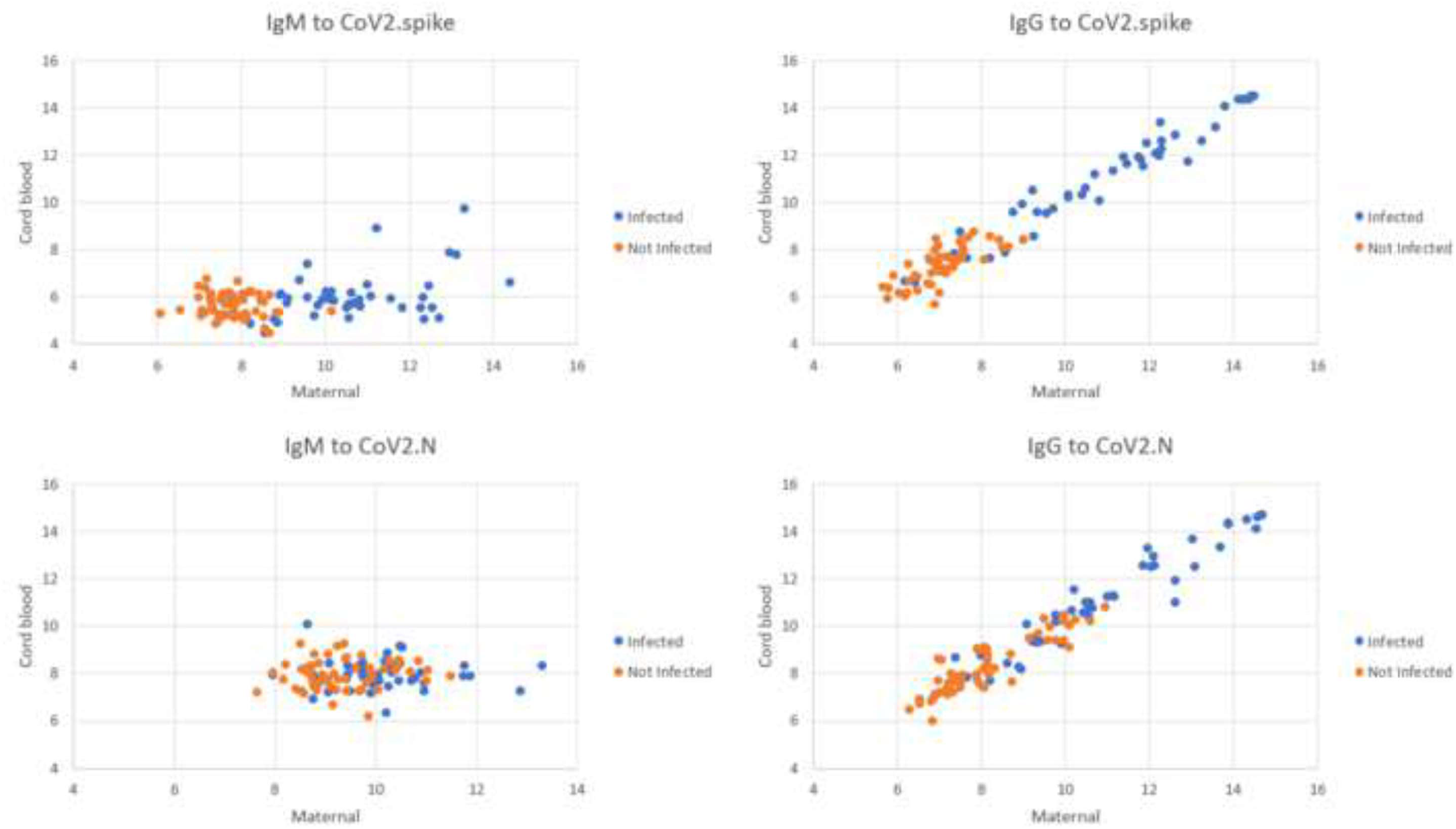
Correlation between maternal and cord blood antibody titers. Maternal and cord blood IgG and IgM to full length spike and nucelocapsid epitope correlation. A linear fit between maternal and cordblood IgG responses shows a slope of 1.01 for both CoV2 spike and CoV2 N antigens.

### SARS CoV-2 Serology and Latency

In a cross-sectional evaluation of examining titers by latency, titers of SARS CoV-2 spike IgG and IgM rise rapidly within the first seven days of infection, while nucleocapsid IgM was similar across time points (Figure 4). In evaluating maternal and cordblood serology in relation to latency, COVID disease>7d from delivery was specifically positively correlated with maternal S-IgG (r=0.42, p=0.01), N-IgG (r=0.47, p=0.001), RBD-IgG (r=0.37, p=0.03), NTD-IgG (r=0.34, p=0.04) and S-IgM (r=0.36, p=0.03) and RBD-IgM (r=0.36, p=0.03), but not N-IgM (r=0.14, p=0.88).

**Figure 4:**
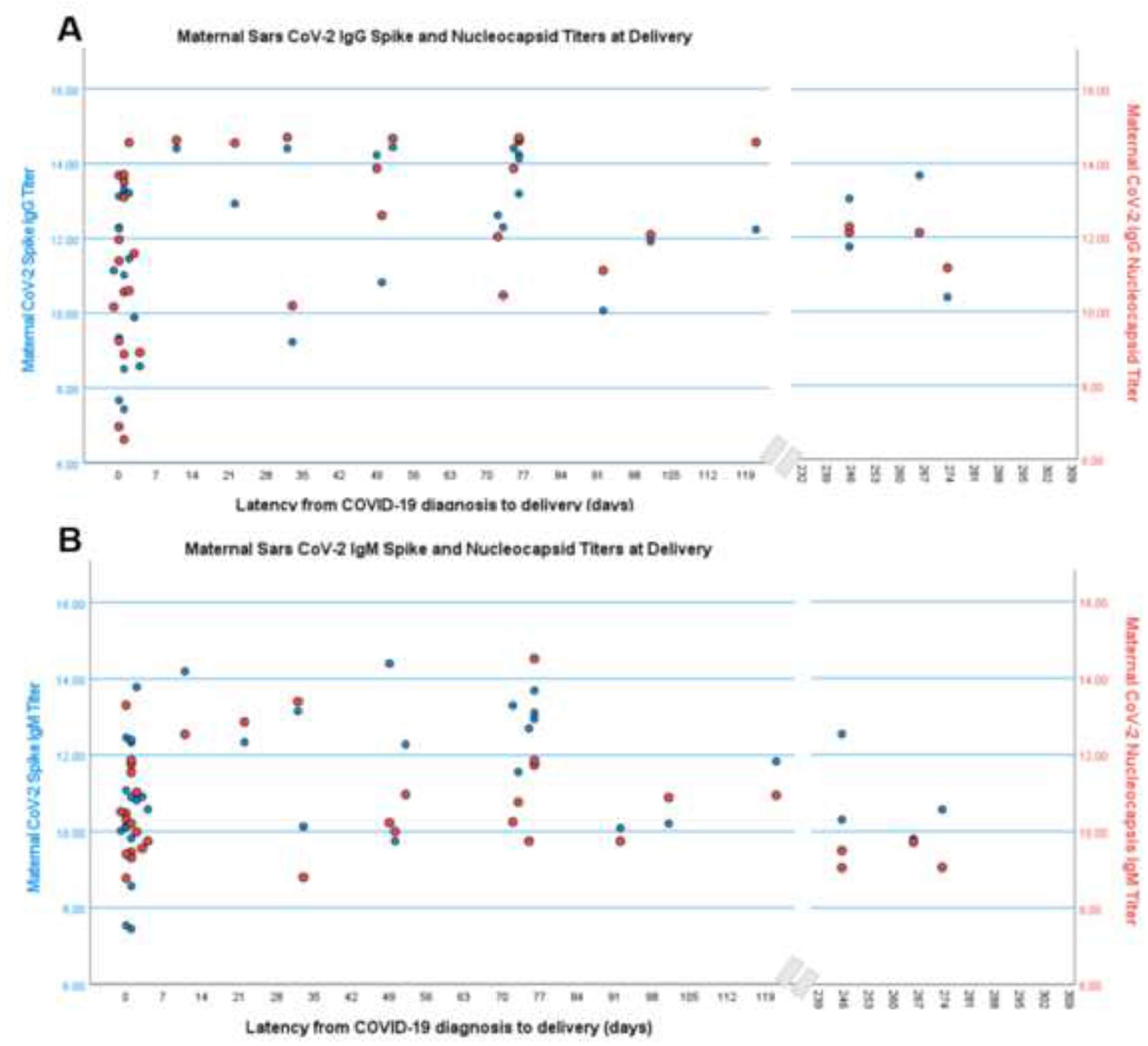
Relationship between latency and maternal antibody titers for IgG and IgM full length spike and nucleocapsid epitopes. Data from N=36 documented COVID-19 infections. Data reported as natural log-transformed luminescence signal.

### SARS-CoV-2 Serology and Placental Pathology

Placental histopathology was available for N=52 participants (N=27 COVID negative and N=25 COVID exposed) and described in Table 2. In looking at antibody titers, maternal vascular malperfusion was significantly correlated with CoV-2 spike IgG (r=0.36, p=0.009), CoV-2 RBD IgG (r=0.32, p=0.02), and CoV-2 N (r=0.32, p=0.02). More correlations existed with CoV-2 IgM epitopes with maternal vascular malperfusion positively correlated with CoV-2 IgM spike (r=0.31, p=0.03), RBD (r=0.29, p=0.04), NTD (r=0.38,p=0.005), and N (r=0.33, p=0.02). CoV-2 IgM N was also positively correlated with placental intervillous thrombosis (r=0.30, p=0.03). Mean maternal N-IgM and S-IgM were significantly higher in those with maternal vascular malperfusion (10.3±1.5 vs 9.6±0.8, p=0.03 and 10.0±2.0 vs 8.8±1.9, p=0.02 respectively), as were N-IgG and S-IgG (10.6±2.7 vs 9.1 ±2.0, p=0.02 and 10.4±2.9 vs 8.3±2.4, p=0.01 respectively) (Figure 5).

**Table 2:** Placental pathological findings associated with COVID-19 exposure.

| Demographics | Maternal COVID PCR<br>and Serology<br>negative<br>(N=27) | Maternal COVID PCR<br>or Serology positive<br>(N=25) | P-value |
| --- | --- | --- | --- |
| Placenta unremarkable | 6 (22) | 1 (4) | 0.10 |
| Acute inflammatory pathology | 7 (26) | 9 (36) | 0.43 |
| Chronic inflammatory pathology | 1 (4) | 0 (0) | 0.33 |
| Maternal vascular malperfusion | 8 (30) | 14 (56) | 0.05 |
| Fetal vascular malperfusion | 1 (4) | 0 (0) | 0.33 |
| Intervillous thrombus | 3 (11) | 3 (12) | 0.92 |

**Figure 5:**
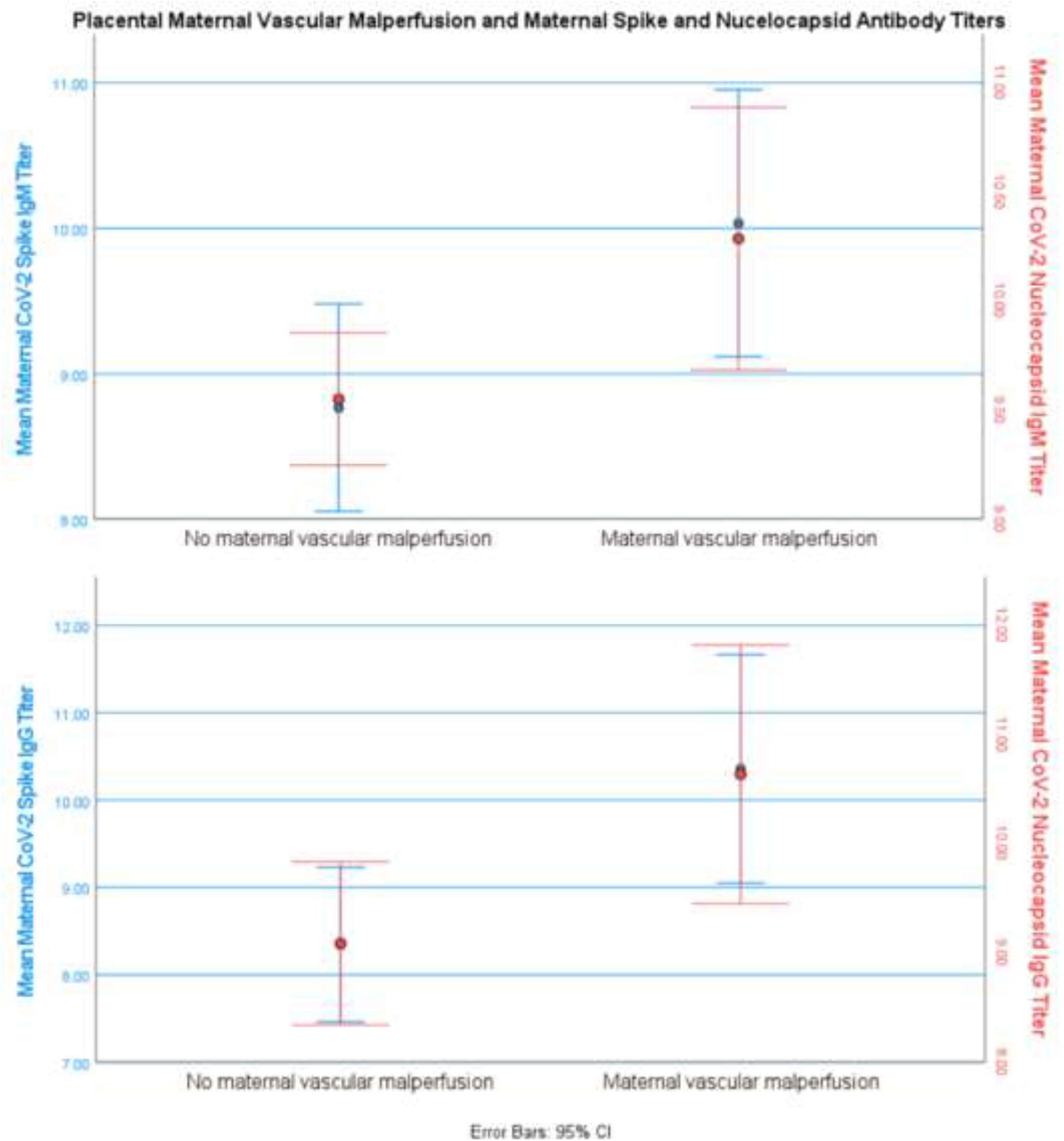
Relationship between maternal spike and nucleocapsid antibody titers and presence of placental maternal vascular malperfusion. Data from N=22 with maternal vascular malperfusion documented on placental histopathology and N=30 with no evidence of maternal vascular malperfusion. Data reported as natural log-transformed luminescence signal. Mean maternal N-IgM and S-IgM were significantly higher in those with maternal vascular malperfusion (10.3±1.5 vs 9.6±0.8, p=0.03 and 10.0±2.0 vs 8.8±1.9, p=0.02 respectively), as were N-IgG and S-IgG (10.6±2.7 vs 9.1 ±2.0, p=0.02 and 10.4±2.9 vs 8.3±2.4, p=0.01 respectively)

## Discussion

### Principle Findings

There is highly efficient transfer in maternal to cordblood IgG, with IgG response to nucleocapsid and spike and IgM to spike (full length) showing the highest specificity. IgG antibodies are cross reactive with related CoV-1 and MERS spike epitopes while IgM, which largely does not cross the placenta, is highly SARS Co-V-2 specific. Our results suggest: 1) Both nucleocapsid and full length spike IgG and IgM antibody epitopes should be included in evaluating serologic response in future studies of disease or vaccine development 2) Serological profile functions as a proxy for latency from disease exposure and in the absence of known disease (i.e. lack of access to testing) and 3) Cord blood, which is effectively depleted of the highly specific maternal IgM response, may have significantly different fine-specificity than maternal blood, despite the high efficiency of IgG transfer.

### Results in the Context of What is Known

This study adds to existing studies through a comprehensive evaluation of maternal and cordblood serological response. One study including 83 mother/baby dyads examined only spike-RBD and IgG and IgM and similarly identified strong correlation between mother and cordblood as well as latency to delivery^19^. Another study of 88 mother/baby dyads identified that IgM peaked at 15 days while IgG peaked at 30 days, which is consistent with our findings regarding maternal and cordblood N-IgG titers and latency >30 days from delivery, although that testing platform was semi-quantitative using a combination of S-thus did not provide granular detail on antibody epitopes and specificity^18^. A third study^22^ looked at 63 mother/baby pairs and examined anti-RBD and anti-N IgG. The limited positive serology reported (65-70% of PCR+ mothers) demonstrates the limitations of examining isolated antibody epitopes, and specifically of RBD only epitope.

### SARS CoV-2 Immunity

While prior studies have focused on SARS-CoV-2 spike protein, we evaluated a range of epitopes. Our results demonstrated high sensitivity of our platform with detection of spike (full length)- IgM and IgG in ∼90% of documented PCR infection. We found that, as in non-pregnant patients, high levels of N-IgG and IgM titers in those with a history of COVID-19^23^. Studies in non-pregnant individuals identified IgM peaks in N and S epitopes in second week of infection^24^. We found that maternal nucleocapsid antibodies demonstrated the highest specificity for documented COVID-19 infection and were highly expressed in both maternal and cordblood samples. Nucleocapsid proteins of many coronaviruses are highly immunogenic and highly expressed during acute infection^25^. While the focus of vaccine and monoclonal antibody therapeutics have been on spike antigen, these results, consistent with other studies in non-pregnant adults, highlights the potential import of N-antibodies in SARS-CoV-2 immunity and target for therapy. Finally, similar to the reports cited above^18,19,22^, we found a high degree of correlation and a linear fit with a slope of 1.0, between maternal and cordblood IgG with cordblood IgG concentrations, indicating highly efficient transfer of CoV-2 IgG antibodies. Finally, in examining serology across latency we found that antibody titers rise rapidly after 7 days and persist >100 days is associated with increased antibody responses, specifically against nucleocapsid and spike epitopes demonstrating these antibody responses appear durable rather than short lived.

Unique to our study, we also evaluated cross reactivity to related viruses by comparing baseline (COVID-19 negative) antibody response against related coronavirus spike antigen to the antibody response in those with COVID-19. We found that while maternal IgG antibodies were cross reactive with related MERS and CoV-1 spike epitopes, IgM was highly specific for CoV-2 and had similar response to MERS and CoV-1 as those without COVID-19. Given that placental antibody transfer is IgG limited, this suggest that cord blood antibody responses may be more cross-reactive and lack some of the CoV2-specific responses found in the IgM of maternal blood. Previous work on adult patients from South Korea indicate that IgM and IgG have a distinct antibody profiles^20^. There is also indication that the profile of the IgM vs IgG response is indicative of functional activities and subsequent disease severity^26^. Cross-reactivity between the different CoVs has been a subject of debate. As the antibody profiles of IgM and IgG differ and are associated with distinct biological functions, it is likely that the lack of IgM in the cord blood may fail to transfer at least some of the protective immunity from the mother to the child. This is further highlighted by recent work demonstrating the importance of IgM in SARS-CoV-2 neutralizing activity in adults^27,28^ and another study comparing pediatric and adult responses which found pediatric response was predominantly anti-spike IgG in contrast to adult IgG, IgM, and IgA against both spike and nucleocapsid epitopes, and had reduced neutralizing activity compared to adults^29^. These systematic differences in isotype and epitope specificity raise the possibility that cordblood, which is effectively depleted of the highly specific maternal IgM response, may have significantly different fine-specificity and may therefore have different neutralizing activity than maternal blood, despite the high efficiency of IgG transfer.

### SARS CoV Serology and Perinatal Outcomes

As in prior studies^30^, we have identified an increased rate of preterm in the setting of COVID-19 exposure, even in this small cohort. We also identified a positive correlation with higher SARS CoV-2 antibody titers and placental maternal vascular malperfusion. This placental finding with COVID-19 has been previously reported^31,32^, and our finding of higher titers related to this finding rather than just COVID exposure, suggest that time from illness, and potentially chronic or downstream effects from COVID-19 lead to placental pathology rather than the acute infectious state (ie within 7 days from infection).

### Strengths and Limitations

This study has a number of strengths. Other evaluations of COVID-19 serology in the mother/baby dyad focused on limited antibody epitopes. We have provided a comprehensive evaluation of maternal and neonatal cord blood serological response looking across the array of SARS CoV-2 antibody epitopes and specificity of response through evaluation of cross reactivity to related viruses. This is a unique examination of serological profile not only in the mother, but also in what is passively acquired by the neonate, and the implications for passive immunity. Although a single institution, our population is diverse, improving external validity.

This study has limitations as well. The number of mother/baby dyads limited our ability to provide a more detailed evaluation of serological changes over time. There may be antibody epitope correlations we were not powered to detect. Finally, while we identified differences in maternal and cord blood antibody signature that could impact immunity, we did not evaluate functional activity.

## Conclusion

Maternal COVID-19 exposure is associated with specific maternal and cord blood antibody signature, with nucleocapsid and full length spike epitopes demonstrating highest specificity in distinguishing exposed vs non-exposed individuals. Serologic profile relates to latency from exposure. There is highly efficient transfer in maternal to cord blood IgG antibodies. IgG antibodies are cross reactive with related CoV-1 and MERS spike epitopes while IgM, which cannot cross placenta to provide neonatal passive immunity, is highly SARS CoV-2 specific, suggesting there may be important qualitative distinctions between maternal immunity and neonatal passive immunity.

## Data Availability

All data produced in the present study are available upon reasonable request to the authors

## Acknowledgements

We would like to acknowledge our senior research coordinator Brandy Firman as well as the Thomas Jefferson University Hospital Labor and Delivery teams who assisted in sample collection. We would like to thank Ms. Jessica Bolton (WRAIR) for technical assistance in the serological analysis. Collection of samples was funded in part through NIH grant 3R21HD101127-01S1 (PI RCB) and a Pilot Grant (ZA) through an Institutional Development Award (IDeA) from the National Institute of General Medical Sciences of the National Institutes of Health under grant number U54-GM104941 (PI: Hicks). RCB is supported by PhRMA Faculty Development Award.

## Disclaimer

Material has been reviewed by the Walter Reed Army Institute of Research. There is no objection to its presentation and/or publication. The opinions or assertions contained herein are the private views of the authors, and are not to be construed as official, or as reflecting the views of the Department of the Army or the Department of Defense. The investigators have adhered to the policies for protection of human subjects as prescribed in AR 70-25. This paper has been approved for public release with unlimited distribution.

